# Faecal cytokine levels of preterm infants coupled with microbiome profiles represent a potential non-invasive method to predict severity of necrotizing enterocolitis

**DOI:** 10.1101/2022.10.24.22281217

**Authors:** Christian Zenner, Lisa Chalklen, Helena Adjei, Matthew J. Dalby, Suparna Mitra, Emma Cornwell, Alex Shaw, Kathleen Sim, J. Simon Kroll, Lindsay J. Hall

## Abstract

**Objectives:** Necrotizing enterocolitis (NEC) is a life-threatening disease, and the most common gastrointestinal emergency in premature infants. Accurate early diagnosis is challenging. Modified Bell’s staging is routinely used to guide diagnosis, but early diagnostic signs are non-specific, potentially leading to unobserved disease progression, which is problematic given the often rapid deterioration observed in NEC infants. New techniques, using biomarkers as diagnostic tool to improve diagnosis of NEC, are emerging. Here we investigated faecal cytokine levels, coupled with gut microbiota profiles, as a non-invasive method to discover specific NEC-associated signatures that can be applied as potential diagnostic markers.

**Study design:** Premature babies born below 32 weeks of gestation were admitted to the 2-site neonatal intensive care unit (NICU) of Imperial College hospitals (St. Mary’s or Queen Charlotte’s & Chelsea) between January 2011 and December 2012. All but two babies received a first course of antibiotics from birth onwards. Faecal samples from diapers were collected consecutively during the NICU stay.

**Results:** Evaluation of microbiota profiles between the study groups revealed only minor differences. However, at later time points, significant changes in microbiota structure were observed for Firmicutes, with *Enterococcus* being the least abundant in Bell stage 2/3 NEC. Faecal cytokine levels were similar to those found in previous studies evaluating systemic cytokine concentrations in NEC settings, but measurement in faeces represents a non-invasive method to evaluate the early onset of the disease. For IL-1α, IL-5 and IL-10, a significantly rising gradient of levels were observed from healthy to NEC1 to NEC2/3.

**Conclusions:** Differences in certain faecal cytokine profiles in patients with NEC indicate their potential use as diagnostic biomarkers to facilitate earlier diagnosis. Additionally, associations between microbial and cytokine profiles, contribute to improving knowledge about NEC pathogenesis.

## Introduction

Necrotizing enterocolitis (NEC) is a life-threatening disease that primarily affects very low birthweight (VLBW) preterm infants born weighing less than 1500g (1). The estimated average incidence of NEC cases across 27 studies conducted worldwide is ∼7% among VLBW infants (2). However, contrasting regional differences are reported in the literature, with a prevalence of NEC of 25.4% for enteral fed and low birth weight infants admitted to public hospitals in Addis Ababa, Ethiopia (3), compared to only 1.6% in VLBW infants in Japan (4).

NEC is a clinical diagnosis of exclusion, not directed by macroscopic or microscopic characteristics. Symptoms range from slowly progressing and nonspecific signs to evidence of extensive gastrointestinal disorders. The central component of NEC is an advancing ischaemic necrosis of the terminal ileum and colon, often resulting in bowel perforation (5). Infant mortality is high in diagnosed NEC cases, and even higher in infants requiring surgery to remove sections of necrotic bowel (6).

Although clinical manifestations of the disease have been known since the 1940s (7), its aetiology remains incompletely understood and is often described as multifactorial (8). The most important contributing factors for the development of NEC is prematurity, including low birth weight and low gestational age (9, 10). Other potential factors are formula feeding (11), prolonged parenteral feeding (12), and an abnormal microbial colonization (13), potentially leading to a perturbed state in the premature intestine (14, 15). The gut of vaginally delivered and breast-fed term babies is typically dominated by bacteria of the genus *Bifidobacterium*, whereas preterm infants, who are often born by caesarean section and receive antibiotic treatment, are populated by genera such as *Enterococcus, Klebsiella*, and *Escherichia*.

Overgrowth of these potentially pathogenic bacteria within the gut microbiota, and/or colonisation of the preterm gut by hospital-acquired pathogens plays a crucial role in the onset of NEC. Frequently detected bacteria occurring in association with NEC include *Clostridium spp*., *Enterococcus spp*., *Escherichia coli, Pseudomonas aeruginosa, Salmonella spp*., *Klebsiella spp*., *and Staphylococcus spp*. (16). These potential pathogens can be partially suppressed by supplementation with probiotics including *Bifidobacterium* spp., which is also associated with a 50% reduction in NEC incidence (17, 18).

More recently, genetic predispositions such as single nucleotide polymorphism in potential NEC associated genes have been identified as risk factors (19, 20).

The prognosis for infants diagnosed with NEC is poor, with survivors at risk of long-term neurodevelopmental limitations and growth restrictions (21-23). The Bell staging criteria were introduced in 1978 to classify different stages of illness severity, suggest disease management, and guide treatment (24), and were later refined in 1986 (25). Various other staging criteria for NEC have been proposed by expert neonatologists, including the Vermont Oxford Network definition, Centers for Disease Control and Prevention definition, Gestational Age-Specific Case Definition of NEC, Two of 3 rule, Stanford NEC score and International Neonatal Consortium NEC workgroup definition. However, modified Bell staging remains the most frequently used (26), despite questions remaining about its reliability (27).

Researchers have focused on additional measures including the infant gut microbiome that could better predict cases of NEC. Dobbler et al. reported that both lower microbial diversity and bacteria belonging to the family *Enterobacteriaceae* correlated with NEC, with *Citrobacter koseri* and *Klebsiella pneumoniae* being the most abundant species within this family (28). Low bacterial diversity in combination with high abundance of Proteobacteria prior to the onset or at diagnosis of NEC has been confirmed by other studies (29-35). In contrast, Cassir et al. showed a strong association between *Clostridium butyricum* and NEC incidence and identified cytotoxic activity in the supernatant of cultured C. *butyricum* isolates (36). The role of the gut microbiota in the development of NEC remains complex and is likely to be dependent on NICU location (i.e. circulating nosocomial pathogens) and underlying individual microbial communities present in the preterm infant gut.

Human milk oligosaccharides (HMOs) are now a topic of research interest due to their role in feeding specific bacteria, especially *Bifidobacterium*, which are not typically abundant in the preterm infant gut microbiota (37). Sodhi et al. recently suggested the HMOs 2’-fucosyllactose and 6’-sialylactose protect against the development of NEC through the inhibition of Toll-like receptor (TLR) 4 signalling (38). Masi et al. showed that the concentration of the HMO disialyllacto-N-tetraose (DSLNT) was lower in the breast milk of mothers of NEC infants and associated with a lower abundance of *Bifidobacterium* species (39).

The role of cytokines and pro-inflammatory mediators in NEC have been extensively reviewed. In particular, increased levels of TLR 4, IL-18, IFNγ, Platelet-activating factor (PAF), IL-6, IL-8, IL-1β, and NF-κB have been linked to NEC severity, while deficiencies of TLR 9, IL-1R8, IL1-Ra, TGFβ_2_, PAF-acetylhydrolase, and IL-10 pave the way for NEC-associated inflammation (40).

Novel approaches are needed, to provide guidance to clinicians and healthcare professionals to select the appropriate therapy (41). Previous studies have aimed to find suitable and robust biomarkers that may be used to predict NEC, including platelet counts (42), and levels of C-reactive protein (43), serum amyloid A (44), claudin proteins (45), plasma citrulline (46, 47), endogenous RNA molecules (48), volatile organic compounds (49), and lipocalin-2 and calprotectin (50). Systemic cytokine concentrations have been suggested as potential biomarkers for the prediction of NEC and disease outcome (40, 51-54). Rising cytokine levels were highly specific for the diagnosis of neonatal sepsis, but additional (non-invasively assayed) biomarkers are needed for high specificity and sensitivity to predict NEC (55).

In this study we evaluate the gut microbiota profiles and the measurement of faecal cytokine levels as a rapid and non-invasive tool for the early detection of NEC.

## Methods

### Study design

Samples were provided from a study published in 2015 (15). 16S rRNA gene amplicon sequencing data was re-analysed with updated reference databases and additionally, cytokine profiles were measured in faeces. This exploratory study included infants born <32 weeks of gestation, without severe congenital birth defects. Infants were admitted to the Imperial College Healthcare NHS Trust neonatal intensive care unit (NICU) between January 2011 and December 2012. In total, 39 individuals were included in the study (Bell stage 1: n=7; Bell stage 2/3: n=11; healthy controls: n=21). Probiotics and H2-receptor antagonists were not used within the NICU at the time of recruitment and sampling. Patient IDs were blinded. Only members of this research group had access to patient information.

### Ethics declaration

This work is part of the study ‘Defining the Intestinal Microbiota in Premature Infants’ (NeoM – The Neonatal Microbiota study) (ClinicalTrials.gov Identifier NCT01102738), approved by West London Research Ethics Committee Two, United Kingdom (Reference number: 10/H0711/39). Parents gave written approval for their infants to participate in the study.

### Sample collection

Research nurses collected faecal samples from diapers, stored the material in sterile tubes at 4°C until transportation to the lab. Neonatal consultants confirmed diagnosed NEC cases (Bell stage 2/3 by Bells’ modified staging criteria). Multiple samples were taken from individuals included in the study during their stay in NICU. Sample numbers were as follows: Bell stage 1 NEC n=23; Bell stage 2/3 NEC n=47; healthy controls n=86.

### Cytokine measurement

Faecal samples were homogenized with PBS using a FastPrep® Bead Beater (4.0m/s, 3min), centrifuged (14,000rpm, 15min) and 25µl of supernatant was used for the assay. Samples were analysed using MULTI-SPOT™ plates, MESO Quickplex SQ120 and discovery workbench software according to the manufacturer’s protocol. Pre-coated immunoassays V-PLEX Proinflammatory Panel 1 (human) and V-PLEX Cytokine Panel 1 (human) were used to detect a set of 20 different cytokines: IFNγ, IL-1β, IL-2, IL-4, IL-6, IL-8, IL-10, IL-12p70, IL-13, TNFα, GMCSF, IL-1α, IL-5, IL-7, IL-12p40, IL-15, IL-16, IL-17A, TNFβ, and VEGF-A. If cytokine values drastically exceeded comparable sample values, the sample was excluded from the analysis. Samples not reaching the lower limit of detection were generally considered as very low and were taken into account without statistical resolving.

### DNA extraction

DNA was extracted from faecal samples using the FastDNA™ Spin Kit for Soil (MP Biomedicals), according to the manufacturer’s instructions with a modified elution step with Tris (10mM) low-ethylendiaminetetraacetic acid (0.1mM) buffer. Two homogenization steps were included for 40s at 6.0m/s. The concentration of DNA was assessed using a NanoDrop™.

### 16S rRNA gene amplification of V3-V5 region

The V3-V5 region of the 16S ribosomal RNA gene was amplified for each sample using primers recommended by the NIH Human Microbiome Project (56). The forward primer contained a

454 Life Science primer B sequence and primer 357F (F: 5’ CTATCCCCTGTGTGCCTTGGCAGTCTCAGCCTACGGGAGGCAGCAG 3’). The reverse primer contained a unique 12 bp error-correcting Golay barcode to tag each sample (denoted with ‘N’) and the bacterial 16S primer 926R (R: 5’ CCATCTCATCCCTGCGTGTCTCCGACTCAG-NNNNNNNNNNNN-CCGTCAATTCMTTTRAGT 3’). Each reaction contained 1µl of each forward and reverse primer (10µM), 1µl of template DNA, 0.25µl of 5U/µl FastStart™ High Fidelity DNA Polymerase (Roche), 0.5µl dNTP mix (10mM each), 2.5µl FastStart™ 10x Buffer#2, 11.25µl molecular biology grade water, 1µl of 10g/ml Bovine serum albumin and 6.5µl of 5M betaine. Thermal cycling was performed as follows: 2min 94°C, followed by 35 cycles of denaturation (20s at 94°C), annealing (30s at 50°C) and extension (5min at 72°C).

### Amplicon Clean-Up, Size selection, quantification, and sequencing

PCR products were cleaned using the Agencourt AMPure XP system according to the manufacturer’s instructions. Amplicons of the correct size were selected using 2% E-Gel SizeSelect gels (Invitrogen). Purified amplicons were quantified in duplicate using the Quant-iT PicoGreen dsDNA assay kit on a FLUOstar Omega microplate reader. Amplicons were pooled in equimolar concentrations. 16S rRNA gene amplicon sequencing was performed on a 454 Life Sciences GS FLX Titanium machine following the Roche Amplicon Lib-L protocol.

### 16S rRNA sequencing data analysis

Roche 454 pyrosequencing data in standard flowgram format was transcribed to fastq format using Bio.SeqIO.SffIO module in biopython. Single fastq files were remultiplexed using the perl script remultiplexor (available at https://www.imngs.org). Remultiplexed sequencing data was processed with the integrated microbial NGS platform (IMNGS) (57) with parameters set as follows: Barcode mismatches, 1; quality trim score, 10; min. read length 100bp; max. read length 1000bp; max. rate of expected error, 2% of sequence length; min. alignment id 70%. Operational taxonomic units (OTUs) were clustered at 97% sequence similarity, using a cutoff of ≥0.25% relative abundance in at least one sample. Data was further analysed and visualized using RHEA (58), a modular pipeline for microbial profiling of 16S rRNA gene amplicon data, using R(v4.0.5) and Rstudio (v1.4.1106). Samples not achieving specific QC criteria (>1000 reads/sample; rarefaction curves Suppl. Fig 1) were excluded from the analysis, leading to reduced sample numbers: Bell 1 NEC 1 n=18; Bell 2 NEC 2/3 n=41; healthy controls n=63.

**Fig1:**
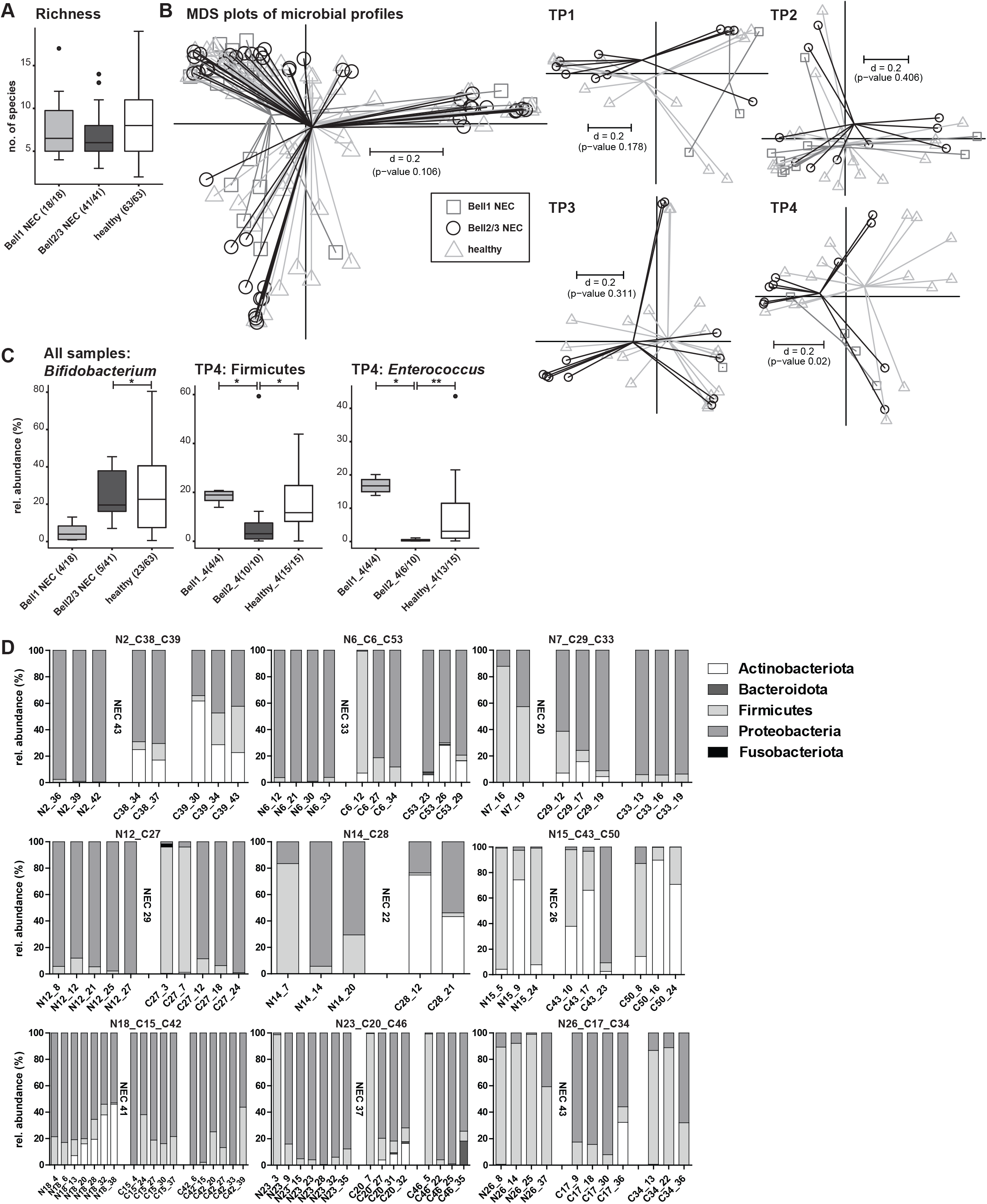
A: Alpha-diversity shown as richness. B: Inter-sample differences shown as multi-dimensional scaling plots based on generalized Unifrac distances across all samples and separated by different timepoints. C: Taxonomic differences across all samples (Bifidobacterium) and at timepoint 4. Numbers in brackets indicate the number of samples positive for the observation. D: Over time age matched taxonomic profiles at the phylum level of preterm babies that developed NEC (left) compared to healthy individuals (right). The DOL of NEC diagnosis is indicated after NEC samples. P-value summary: *<0.05; **<0.01.

### Statistical testing

Cytokine profiles were evaluated pairwise between groups using Mann-Whitney-U Test. The following methods were applied for 16S rRNA gene amplicon data: Fishers Exact Test, Wilcoxon Rank Sum, and Kruskal-Wallis Rank Sum Test. The method used is referenced in the respective paragraph or figure. Multidimensional scaling plots are based on generalized UniFrac distances. The p-value was calculated using PERMANOVA.

## Results

A total of 39 preterm infants with a gestational age <32 weeks were included in this study, 7 were diagnosed with Bell stage 1 NEC, 11 were diagnosed with Bell stage 2/3 NEC and 21 were healthy controls (not diagnosed with NEC). Detailed information about participants and sample numbers are represented in table 1. All but two babies received a first course of antibiotics from birth onwards. Faecal samples from diapers were collected longitudinally during their NICU stay.

**Table 1:**
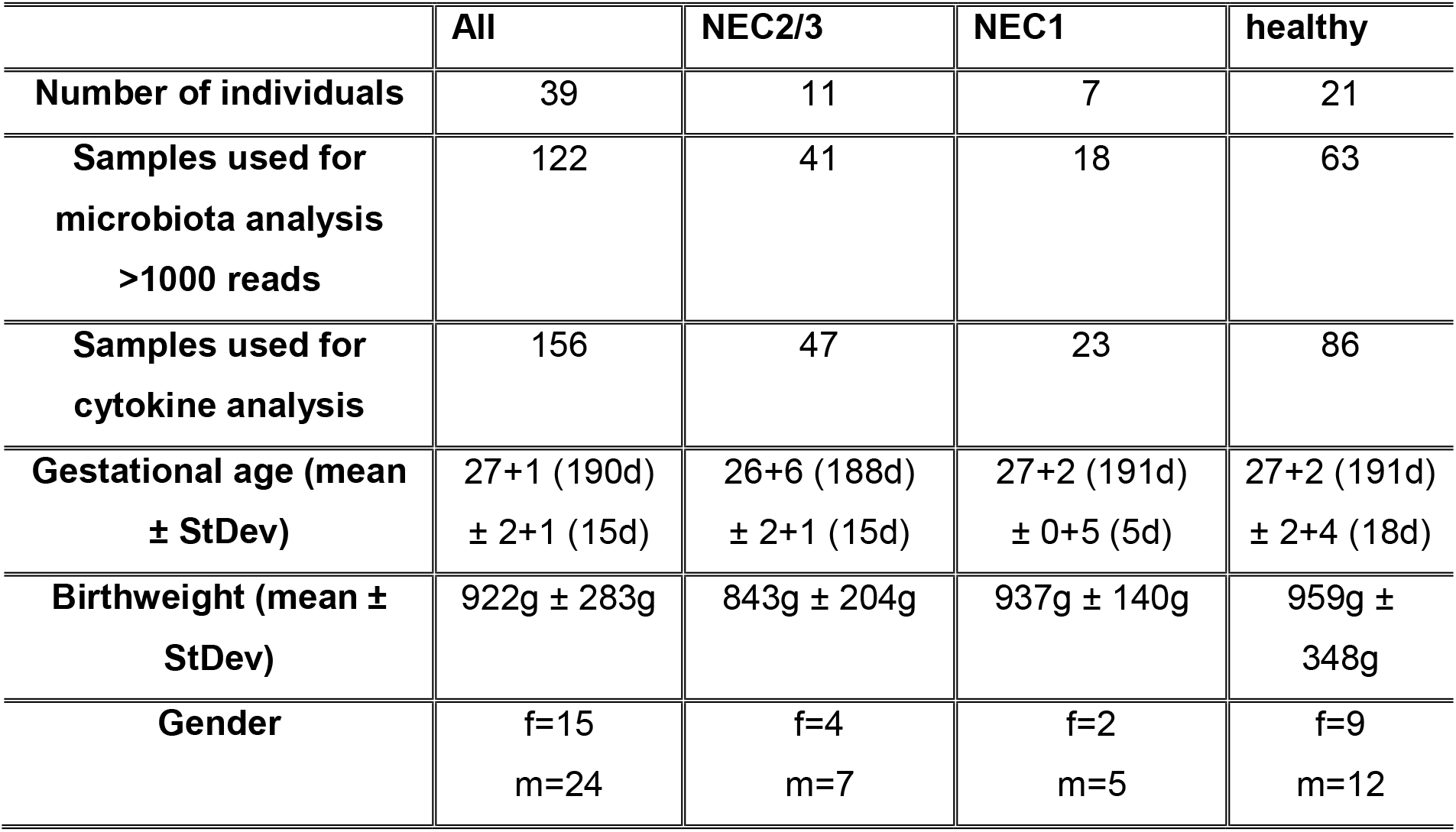
Cohort information of study participants

Characterisation of the neonatal gut microbiome of these preterm infants was carried out using 16S rRNA gene amplicon sequencing. An average of 7.8 (±3.6) OTUs (a proxy for bacterial species) was detected across the three infant groups. Healthy infants contained a mean of 8.4 OTUs/sample, which was lower at 7.6 OTUs/sample in the NEC1 infants and 6.9 OTUs/sample in the NEC2/3 infants, but the differences were not statistically significant (Fig. 1A). The multi-dimensional-scaling (MDS) plot of microbial profiles representing *beta*-diversity showed no significant differences across the three study groups (p=0.106) (Fig. 1B). To detect age-dependant differences, samples were split up into four different time points (TP1: 0-10 days of life (DOL), TP2: 11-20 DOL, TP3: 21-30 DOL, TP4: 31-Maximum age). Significant differences in the *beta*-diversity were detected at time point 4 in the MDS plot (p=0.02) (Fig. 1B). By comparing the groups at taxonomic levels, the only detected significant differences were between Bell stage 2/3 and healthy controls for the order Bifidobacteriales, including family Bifidobacteriaceae and genus *Bifidobacterium* (adj. p=0.0204 for all three taxonomic levels, Fisher’s exact Test, pairwise comparison) (Fig. 1C). At all taxonomic levels, no significant differences were detected at TP1 and TP2. At TP3, a significantly higher relative abundance of *Escherichia-Shigella* in Bell 2/3 was detected compared to the healthy group (p=0.0003, Wilcoxon Rank Sum Test, pairwise, data not shown). At TP4, the microbiota profiles became more clearly different. The phylum Firmicutes was lower in Bell 2/3 (mean rel. abundance 10.0 %) compared to Bell 1 (mean rel. abundance 18.1%) and healthy (mean rel. abundance 15.6%) (adj. p=0.0396, Wilcoxon Rank Sum Test, pairwise comparison, equal p-value for both comparisons) (Fig. 1C). Differences in Firmicutes were mostly represented by differences in the family Enterococcaceae and the subordinate genus *Enterococcus* (NEC1 vs. NEC2/3 adj. p=0.0142; NEC2/3 vs. healthy adj. p=0.0096, Wilcoxon Rank Sum Test, pairwise, values are equal for family and genus) (Fig. 1C). Individuals that developed NEC were further compared with age-matched healthy preterm babies, with phylum profiles measured longitudinally until NEC diagnosis. Only two NEC babies displayed high Actinobacteriota abundance (N15, N18), whilst this phylum was better represented in the healthy control babies. Bacteroidota was generally underrepresented in the studied individuals. Fusobacteria were also rare, and only found in one control baby at one time point (C27_3) (Fig. 1D).

Faecal cytokine concentrations were then analysed to determine differences in these host-associated immune factors. Pro- and anti-inflammatory cytokines play an important role in the development and progression of NEC and systemic levels are often measured. As NEC is primarily an intestinal disease, cytokine concentrations measured in faeces could be more representative of immune activation in NEC.

In these infants almost all measured cytokine concentrations were significantly higher in the NEC 2/3 group (Fig. 2A). Significant differences between NEC1 and NEC2/3 as well as between NEC2/3 and healthy were observed for IL-2, IL-6, IL-10, IL-12p70, IL-12_IL-23p40, IL-13, IL-17A, and Interferon γ (p≤0.0001, Mann-Whitney-U Test) (Fig. 2A). Significantly higher concentrations in NEC1 compared to healthy were observed for IL-1α (p=0.045), IL-10 (p=0.032), and IL-5 (p=0.0074), suggesting that these could be potential markers for the onset and development of NEC. The concentration of these cytokines was further investigated at each time point (Fig. 2B). For IL-1α, significant differences were detected at TP1 (NEC1 vs. NEC2/3, p=0.0307, and healthy vs. NEC2/3, p=0.0177) and TP4 (healthy vs NEC1, p=0.0057, and healthy vs. NEC2/3, p=0.001). For IL-5, significant differences were observed at TP1 (NEC1 vs. NEC2/3, p=0.0106, and healthy vs. NEC2/3, p=0.0004), TP2 (healthy vs. NEC1, p=0.0115, and healthy vs. NEC2/3, p=0.004), TP3 (healthy vs. NEC2/3, p=0.0228), and TP4 (healthy vs. NEC2/3, p=0.0432). Significantly higher levels of IL-10 were found in the NEC2/3 group at all time points, TP1 (NEC1 vs. NEC2/3, p=0.0045, healthy vs. NEC1, p=0.031, healthy vs. NEC2/3, p<0.0001), TP2 (NEC1 vs. NEC2/3, p<0.0001, healthy vs. NEC2/3, p<0.0001), TP3 (healthy vs. NEC2/3, p<0.0001), and TP4 (NEC1 vs. NEC2/3, p=0.0275, healthy vs. NEC2/3, p=0.0003).

**Fig2:**
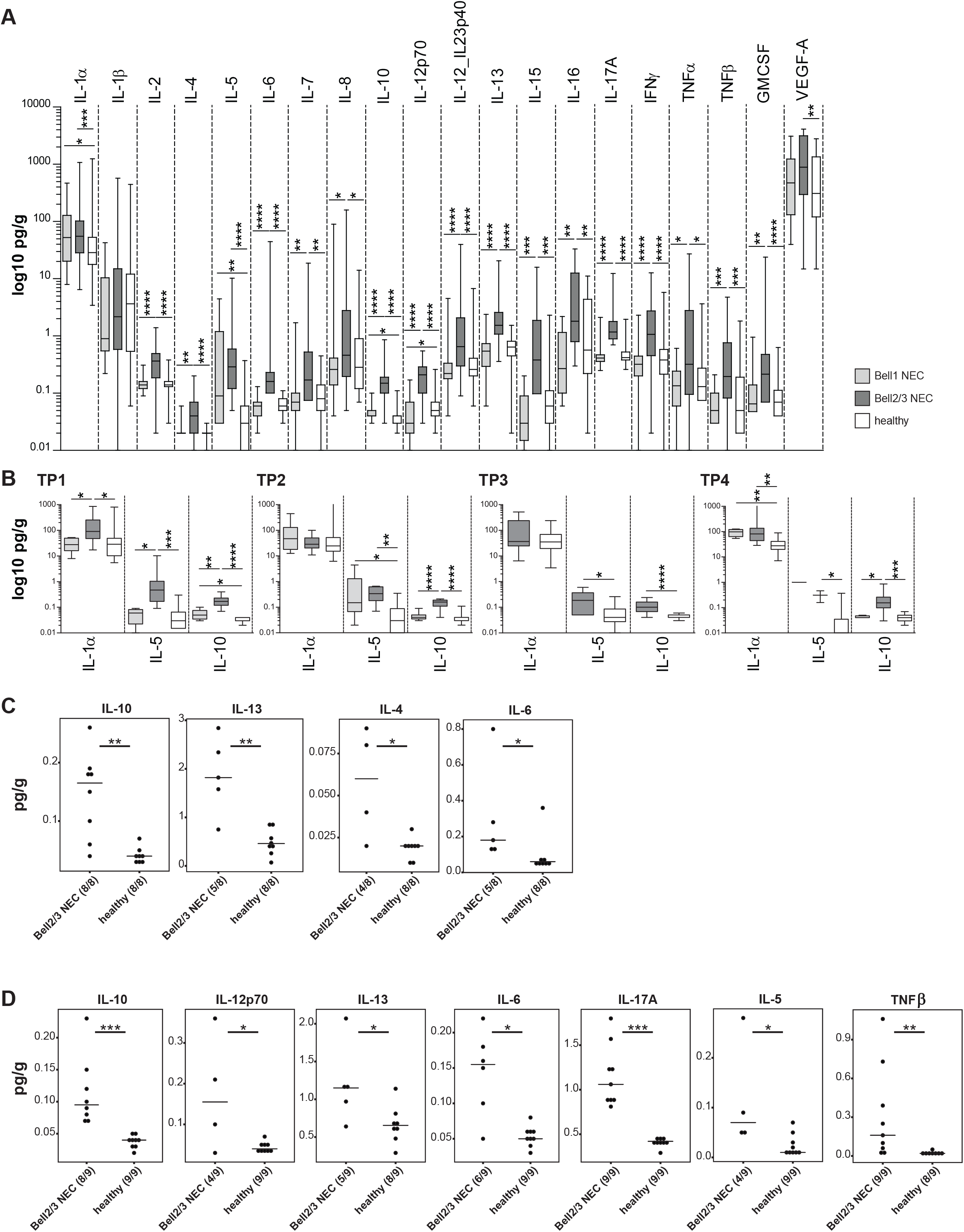
Cytokine levels measured in faecal samples of preterm infants in the three study groups: Bell stage 1 NEC, Bell stage 2/3 NEC, and healthy. A: Across all time points. B: Divided by time points for IL-1α, IL-5, and IL-10. At TP3, as only one sample was present in the NEC 1 group, it was excluded from the analysis. Concentrations in pg/g are plotted on a log 10 scale for better visibility. C: Significant cytokines 5-10 days before NEC diagnosis compared to age matched controls. D: Significant cytokines 11-17 days before NEC diagnosis compared to age matched controls. Numbers in brackets indicate the number of samples positive for the observation. A+B comparisons were statistically analysed with Mann-Whitney-U Test. C+D comparisons were statistically analysed with Wilcoxon Rank Sum Test. P-value summary: *<0.05; **<0.01; ***<0.001; ****<0.0001.

Cytokine profiles were further analysed 5-10 days before the date that NEC was diagnosed and compared with age matched healthy preterm infants (+/- 1 day difference). Significantly higher levels of IL-10 (p=0.0013), IL-13 (p=0.0062), IL-4 (p=0.0293), and IL-6 (p=0.0322) were measured in the Bell stage 2/3 group compared to healthy controls (Fig. 2C). The same analysis was performed 11-17 days before NEC diagnosis with significant differences again detected for IL-10 (p=0.0004), IL-13 (p=0.0335) and IL-6 (p=0.0122), with additional cytokines IL-12p70 (p=0.0294), IL-17A (p=0.0004), IL-5 (p=0.0294), and TNFβ (p=0.0066) also differentiating between NEC and healthy controls (Fig.2D).

## Discussion

Although known for decades, NEC remains a major challenge for neonatologists, given the abrupt onset and rapid progression of the disease. Targeted treatments are still lacking, leading to high mortality rates and leaving survivors with severe long-term disabilities. Prompt timing of treatment is crucial to maximise the chance of survival. In this study, we investigated the preterm infant gut microbiome in combination with faecal cytokine levels to shed light on disease progression. The preterm intestinal microbiota differs greatly from that of term infants: the number of species present is reduced, patterns of colonization are disrupted and the abundance of pathogenic bacteria is increased (59-61). Many studies have reported that reduced gut bacterial diversity is a risk factor for the onset of NEC (28, 30, 33, 62). In our study, samples from the NEC 2/3 group contained the lowest number of OTUs per sample (mean of 6.9), but compared to the other study groups differences were minor and not significant. In terms of taxonomic differences, an enrichment of Proteobacteria and a reduction of Firmicutes and Bacteroidota has often been associated with NEC development (14, 63). However, this was not observed in our study results, with similarly high levels of Proteobacteria found in all study groups. We do detect significantly lower levels of Firmicutes in NEC 2/3 infants at time point (TP4), representing higher Proteobacteria levels, but this was only the case for infants older than 31 days and was not associated with NEC. A variety of reasons could account for differences between studies, including sampling technique, DNA extraction protocols, selection of 16S variable regions, sequencing technique, bioinformatics pipelines, and databases used (64, 65), making comparisons between studies difficult. As numerous bacteria are potentially associated with NEC, i.e. *Clostridium* spp., *Enterococcus* spp., *Escherichia coli, Pseudomonas aeruginosa, Salmonella* spp., *Klebsiella* spp., *and Staphylococcus* spp. (16), a single bacterial signature is not expected. On the other hand, supplementation of probiotic *Bifidobacterium* and *Lactobacillus* is associated with lower abundance of common pathobionts in the preterm gut (17), which is associated with significantly reduced rates of NEC and late onset sepsis (18). Although substantial differences in microbiota profiles were not found in this study between NEC infants and healthy controls, the impact of the microbiome on the immune system including signalling molecules such as cytokines is well known (66). Therefore, the evaluation of faecal cytokine levels is a key aspect of this study. Interestingly, except for IL-1β the faecal concentrations of all measured cytokines were significantly higher in the NEC 2/3 group compared to healthy controls. IL-1s (including IL-1α and IL-1β) are pro-inflammatory cytokines, produced by a variety of cell types, that also induce inflammatory reactions such as tissue damage and fever (67). IL-1 receptor binding triggers the activation of pro-inflammatory transcription factors such as NF-κB and AP-1, which can further induce the production of IL-6, Tumour necrosis factor (TNF) and IL-1 itself (67). Studies on human IL-1α and IL-1β in NEC setting are rare. One study by Benkoe et al. could not identify differences in systemic IL-1β levels in NEC babies compared to healthy controls (51), concordant with the results of our study. For IL-1α, we could identify significantly higher levels in NEC2/3 compared to NEC1 and healthy at TP1, and significantly higher levels in NEC2/3 and NEC1 compared to healthy at TP1 (Fig. 2B). Another study by Ng et al. showed increased systemic concentrations of IL-2, IL-4, IL-6, IL-10, IFNγ, and TNFα in neonatal septicaemia, also including NEC cases (68), corresponding with the results presented in this study for faecal cytokines. We could also show that local IL-10 levels were significantly higher in NEC2/3 compared to NEC1 and healthy at all time points (Fig. 2B). Additionally, the age matched comparison of babies 5-10 or 11-17 days before NEC diagnosis revealed significantly higher levels of IL-10 (Fig. 2D), indicating an induced protective role of IL-10 to counteract inflammation in the gut. This is also supported by high levels of IL-10 in breast milk (69), while low levels of IL-10 in breast milk are correlated with NEC incidence (70). IL-5 primarily promotes activation, survival and adhesion of eosinophils, and is therefore elevated in allergy and parasitosis (71). Interestingly, we observed significantly higher IL-5 concentrations in NEC2/3 at all time points (Fig. 2B), suggesting a hyper-inflammatory state with involvement of eosinophils, coinciding with a study from 2000 (72). While IL-4 and IL-5 were involved in NEC progression in rats (73), Benkoe et al. demonstrated significantly lower IL-4 and IL-5 concentrations in NEC serum samples compared to healthy controls (51).

These findings suggest that faecal cytokine concentrations could provide additional measures in the diagnosis of NEC. Particularly IL-1α, IL-10 and IL-5, which show a rise from healthy to NEC 1 to NEC2/3 and could potentially be used as accessory markers to the current Bell staging that is routinely performed. As faecal samples are taken from diapers they represent a non-invasive method of sample collection, in contrast to blood samples, potentially informative as early as 2 weeks prior to NEC onset. However, the amount of faeces produced by preterm infants can be small making it difficult to get an adequate sample for analysis, particularly in the first days after birth. Additionally, defecation might be reduced further if the gut is already inflamed and the infant is suffering from intestinal symptoms. The timing of sampling and a rapid analysis yielding results within 24h would be essential for the most effective use of faecal cytokine measurement in aiding the diagnosis of NEC. Although it is not common for preterm infants to develop NEC within the first 10 days, one preterm study infant was diagnosed with NEC in that time period, whilst the other NEC cases were diagnosed between 20 and 43 days old. Our data indicates that profiling faecal cytokine levels, particularly IL-5 and IL-10, from 14 days onwards, and regular testing every third day for increasing levels could act as a predictive test, warning of developing NEC. However, robust reference values of healthy preterm infants and other NEC cases from other NICUs will be required to define highly selective and sensitive cytokine thresholds, in order to provide additional information and guidance to neonatologists in the diagnosis of NEC.

Additional research will also need to test and validate different platforms for faecal cytokine analysis, and compare different preterm infant cohorts to explore cytokine profile variation across different NICUs as a robust markers would be key for next stage studies. Although further testing is required, development of an early diagnosis could refine therapeutic measures, mitigate disease outcomes, increase survival rates and reduce long-term consequences for survivors.

## Data Availability

Data availability:
16S rRNA gene amplicon data is available under BioProject accession number PRJNA889687. Cytokine data is provided in Supplementary Table 1.

## Abbreviations

NEC: Necrotizing enterocolitis
NICU: neonatal intensive care unit
VLBW: very low birthweight
DOL: day of life
OTU: operational taxonomic unit

## Acknowledgement

We thank the participants and their families for their contribution to this study. LJH is supported by Wellcome Trust Investigator Awards (100974/C/13/Z and 220876/Z/20/Z); the Biotechnology and Biological Sciences Research Council (BBSRC), Institute Strategic Programme Gut Microbes and Health (BB/R012490/1), and its constituent projects BBS/E/F/000PR10353 and BBS/E/F/000PR10356. Work at Imperial College was supported by a programme grant from the Winnicott Foundation to JSK, and the National Institute for Health Research (NIHR) Biomedical Research Centre based at Imperial Healthcare NHS Trust and Imperial College London. KS was funded by an NIHR Doctoral Research Fellowship [NIHR-DRF-2011-04-128]. The funders had no role in study design, data collection and analysis, decision to publish, or preparation of the manuscript.

## Data availability

16S rRNA gene amplicon data is available under BioProject accession number PRJNA889687. Cytokine data is provided in Supplementary Table 1.

