## Supplementary figures and images for "Faecal cytokine levels of preterm infants coupled with microbiome profiles represent a potential non-invasive method to predict severity of necrotizing enterocolitis"

### Supplementary Table 1

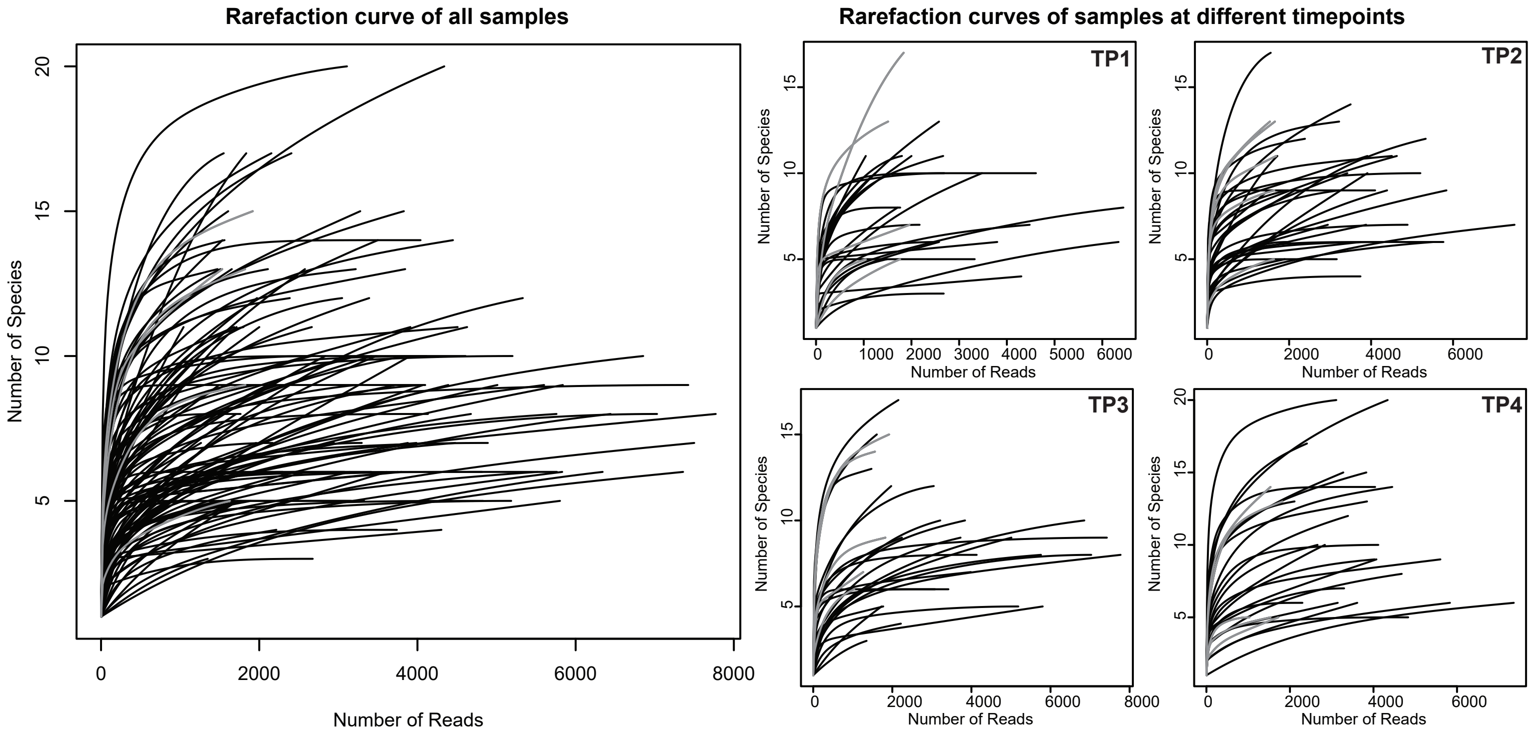
